# Comparison of Accelerated Resolution Therapy (ART) for Post-Traumatic Stress Disorder (PTSD) Between Veterans With and Without Prior PTSD Treatment

**DOI:** 10.1101/2021.04.15.21255572

**Authors:** Tiantian Pang, Lindsay Murn, Dana Williams, Maayan Lawental, Anya Abhayakumar, Kevin E. Kip

## Abstract

**Background:** Post-traumatic stress disorder (PTSD) is a psychiatric disorder commonly caused by a traumatic event(s) and prevalent among service members and veterans. Accelerated Resolution Therapy (ART) is an emerging “mind-body” psychotherapy for PTSD that is generally briefer and less expensive than current first-line treatments, such as cognitive processing therapy (CPT) and prolonged exposure (PE) therapy.

**Objective:** This study aimed to examine the results of ART for treatment of military-related PTSD, with stratification by prior history of PTSD treatment, including refractory PTSD following receipt of guideline-driven first-line psychotherapy.

**Methods:** The study compares the PTSD treatment results of ART between (military service members and/or) veterans with a prior history of PTSD treatment (medication only, n=40; first-line psychotherapy, n=33; other psychotherapy, n=42) and a treatment-naïve group (n=33). Participants were assessed at baseline, post-treatment, and 3- or 6-month follow-up using PCL-M scores (PTSD checklist).

**Results:** Mean age was 43.8 years, 95% male, 84% white race. The treatment completion rate was 72% with a mean of 3.5 treatment sessions. Within-group standardized effect sizes for pre-to-post changes in PTSD scores (PCL-M) were large at 1.11, 1.88, 1.03, and 1.48 for the medication only, first-line psychotherapy, other psychotherapy, and treatment-naïve groups, respectively (p=0.02 for between-group comparison). Similar results were observed for measures of depression and anxiety, and baseline to follow-up results was generally similar.

**Conclusions:** In a brief treatment period, ART appears to result in substantial reductions in symptoms of PTSD among veterans, including those previously treated (unsuccessfully) with first-line psychotherapies endorsed by the U.S. Department of Defense (DoD) and Veterans Affairs (VA). These results suggest that ART be considered as a treatment modality for veterans with refractory PTSD.

**Highlights:** Accelerated Resolution Therapy (ART) is a potentially acceptable psychological intervention for treatment-refractory PTSD.

## Introduction

Trauma is ubiquitous. A traumatic event is an experience that is shocking, scary, dangerous, and/or life-threatening (including but not limited to a natural disaster, serious accident or injury, physical abuse, sexual or intimate partner violence, terroristic attacks, or military or combat experience) (American Psychiatric Association, 2020; National Center for PTSD, 2019; National Institude of Mental Health, 2019). Nearly two-thirds of adults have encountered a traumatic or adverse incident during childhood, and 50-60% of adult men and women have experienced at least one trauma in their lifetime, with between 10-20% developing post-traumatic stress disorder (Centers for Disease Control and Prevention, 2020; National Center for PTSD, 2019; U.S. Department of Veterans Affairs, 2019). Post-traumatic stress disorder (PTSD) is a chronic, disabling, and stress-related psychiatric disorder that may occur after exposure to traumatic events(s) and may result in persistent re-experiencing of details related to the trauma(s), avoidance symptoms, negative alterations in cognitions and mood, and alterations in trauma-related arousal and reactivity (American Psychiatric Association, 2013). PTSD rates are particularly high among U.S. military service members and veterans and are accompanied by an increased risk for other mental health concerns like depression, substance misuse, and even suicide (U.S. Department of Veterans Affairs, 2021). As of 2019, there are 17 million military veterans in the United States with 35.6% from the Vietnam era, 22% from the Gulf War (8/1990-8/2001), 21.7% from the post-911 era (9/2001 or later), 6.6% from the Korean War, and 2.2% from World War II (United States Census Bureau, 2019). In 2017, there were nearly 1.3 million active military personnel and 809,000 reserve forces (Department of Defense, 2017). Among the military personnel who had a recent deployment to Iraq and Afghanistan, the prevalence of PTSD has been estimated at 10-20% (Gates et al., 2012; Kok, et al., 2012; Ramchand et al., 2010). A recent longitudinal cohort study found that at least 50% of U.S. service members and veterans suffered from persistent PTSD symptoms and impairment at three-year follow-up; of those, 71% still met criteria six years later (Armenta et al., 2018). Moreover, approximately 11% of Vietnam War veterans experience persistent PTSD-related impairment (Marmar et al., 2015). The increased risk of PTSD in military personnel is associated with a longer cumulative length of deployments, combat-related exposure, disabling injury/illness, and depression (Armenta et al., 2018; Xue et al., 2015). The large number of military personnel in the U.S. with debilitating PTSD illustrates a paramount interest and need for effective treatments. Untreated and/or inadequately treated PTSD can cause distress and lead to a series of health-related comorbidities and interfere with social, educational, and occupational functioning (Bryan et al., 2015; Levine et al., 2014; National Collaborating Centre for Mental Health (UK), 2005; Pietrzak et al., 2011; Wisco et al., 2014).

The VA/DoD Clinical Practice Guideline recommends the use of individual, manualized, trauma-focused psychotherapy for PTSD that has a primary exposure and/or cognitive restructuring intervention (The Management of Posttraumatic Stress Disorder Work Group, 2017). The most well-established “first-line” treatments for PTSD are Cognitive Process Therapy (CPT), Prolonged Exposure (PE) Therapy, and Eye Movement Desensitization and Reprocessing (EMDR) (American Psychological Association, 2017; Cusack et al., 2016; Hamblen et al., 2019; The Management of Posttraumatic Stress Disorder Work Group, 2017; Watts et al., 2013). Unfortunately, clinical response and treatment completion rates with first-line psychotherapies for PTSD - PE and CPT in particular - have been suboptimal. In brief, a review of first-line psychotherapy for military-related PTSD literature included four randomized controlled trials (RCTs) examining PE and five examining CPT reported that clinical outcomes were heterogeneous, nonresponse rates were high, and the benefit of PE and CPT relative to non– trauma-focused treatments was small (Steenkamp et al., 2015). More recent trials of PTSD treatment have had a greater emphasis on combat exposure instead of sexual trauma, used active comparison groups, and examined active-duty personnel treated in-garrison, rather than only veterans (Foa et al., 2018; Nidich et al., 2018; Rauch et al., 2019; Resick et al., 2017; Resick et al., 2015). Across these trials, all active treatments (PE, CPT, present-centered therapy, sertraline, and transcendental meditation) reduced PTSD symptoms, though nonresponse rates were high, and the types of treatment did not significantly differ in their outcomes (Foa et al., 2018; Nidich et al., 2018; Rauch et al., 2019; Resick et al., 2017; Resick et al., 2015; Steenkamp et al., 2020). Overall, the treatment effect was modest at most, with 50% or less of patients assigned to PE experiencing recovery or improvement, and the diagnosis often retained post-treatment, indicating that symptoms improved yet rarely remitted (Foa et al., 2018; Nidich et al., 2018; Rauch et al., 2019; Resick et al., 2017; Steenkamp et al., 2020). Moreover, trials of PE that included standard length of therapy (e.g., 8 weeks) reported treatment non-completion rates of 25% to 48% (Foa et al., 2018; Nidich et al., 2018; Rauch et al., 2019). Thus, Steenkamp et al. (2020) concluded that current first-line psychotherapies have limited efficacy in treating military-related PTSD and do not outperform non-trauma-focused interventions.

Accelerated Resolution Therapy (ART) is an emerging “mind-body” psychotherapy that is considered an alternative treatment for PTSD. Unlike current first-line psychotherapies, ART is short-term - typically delivered in 1-5 sessions over an approximate two-week timeframe. Furthermore, ART does not require additional therapeutic homework or medication, which reduces patient time (and financial) commitment when compared to the 8-15 sessions recommended for PE, CPT, and EMDR protocols (Kip et al., 2012). ART has shown effective reduction in PTSD symptoms in adult civilian and military populations within an average of four treatment sessions with a completion rate of approximately 90% (Kip et al., 2012; Kip et al., 2015; Kip et al., 2013). In addition, ART does not require verbalization of the traumatic event, which may be particularly beneficial for military personnel who cannot share the confidential and sensitive details of the events (Kip et al., 2013), or for those who are reticent to do so, as is required in the more emotionally-demanding therapies like CPT and PE (Steenkamp et al., 2020). Therefore, the treatment protocol of ART may address some of the limitations of the current first-line psychotherapies. ART has a solid theoretical basis (Kip et al., 2014; Kip & Diamond, 2018) and is a promising approach for consideration as an effective trauma-focused treatment modality to meet the specific needs of military service members and veterans who have either never received treatment and or who have treatment-refractory PTSD (Kip et al., 2019; Kip & Diamond, 2018). Thus, the present analysis examines the results of ART used in the treatment of military-related PTSD, with stratification by prior PTSD treatment history, including refractory PTSD following receipt of first-line recommended psychotherapy.

## Method

Two published studies have been conducted on Accelerated Resolution Therapy (ART) for treatment of post-traumatic stress disorder (PTSD) in U.S. military service members and veterans to serve as the basis for this analysis. The first study was an uncontrolled observational prospective cohort study (*n*=188), with clinical assessment performed at baseline, post-treatment, and 6-month follow-up (Kip et al., 2016). Of the 188 screened participants, 148 met the inclusion criteria and initiated treatment with ART. The second study was a two-group randomized controlled trial (*n*=63; Kip et al., 2013). A total of 57 service members or veterans were eligible and enrolled, of whom, 28 out of 29 participants received the ART intervention, and 22 out of 28 subjects from the control group crossed over to receive ART after three months. In total, 50 participants completed the treatment with ART and were assessed with clinical measurements at baseline and post-treatment, and a subset at 3-month follow-up. Similar inclusion and exclusion criteria were used for enrollment, and the screening instrument and recruitment methods were generally similar between the two previously published studies (Kip et al., 2016; Kip et al., 2013). Both studies were approved by the Institutional Review Board at University of South Florida (USF) and all participants provided written informed consent. The present study combined the two data sets for more in-depth analyses.

Demographic, brief medical history, and prior history of PTSD treatment data were collected at baseline. Both studies used similar outcome measures to assess the pre-to-post changes in symptoms of PTSD and its associated comorbidities: the 17-item military version of the PTSD Checklist (PCL-M) (Blanchard et al., 1996; Weathers et al., 1993), the 20-item Center for Epidemiologic Studies Depression Scale (CES-D) for depression (Radloff, 1977); the 21-item State-Trait Inventory for Cognitive and Somatic Anxiety (STICSA) for anxiety (Grös et al., 2007); the 18-item Brief Symptom Inventory (BSI) for psychological distress (Meachen et al., 2008); and the Pittsburgh Sleep Quality Index (PSQI) for sleep quality (Buysse et al., 1989).

### Definitions for Parallel Previous PTSD Treatment History

For the classification of prior PTSD treatment for analysis, two methods were used. First, classification #1 included only participants from the prospective cohort study (Kip et al., 2016). This study included specific documentation on the prior type(s) of psychotherapy received by service members/veterans for treatment of PTSD. This allowed classification of prior “first-line” evidence-based psychotherapy received, particularly prolonged exposure (PE) and/or cognitive processing therapy (CPT). With this information, U.S. military service members/veterans were grouped based on types of prior history of PTSD treatment, as follows: treatment-naïve (*n*=33), medication only (*n*=40), other types of psychotherapy (*n*=42), or first-line psychotherapy (*n*=33). The last three groups were defined as treatment-refractory because study participants reported receiving prior treatment(s) for PTSD, yet were highly symptomatic at the time of assessment for ART.

For the second classification (classification #2), a larger sample was constructed by pooling data from the prospective cohort study and randomized controlled trial, described above (Kip et al., 2013; Kip et al., 2016), yet without the granularity of knowing the specific types of psychotherapy received among all participants. This resulted in study participants being classified into four groups based on their prior history of PTSD treatment, as follows: treatment-naïve (*n*=50), medication only (*n*=43), psychotherapy (*n*=25), or psychotherapy and medication (*n*=80).

### ART Intervention

For both studies, ART was delivered in 1 to 5 treatment sessions with a typical session duration of 60 to 75 minutes. In brief, each session consisted of two core components of trauma-focused therapy - imaginal exposure (IE) and imaginary rescripting (IR) - facilitated with the use of sets of bilateral (side-to-side) eye movements. The bilateral eye movements were performed throughout the treatment session by having the participant follow the ART clinician’s oscillating hand, silently moving their eyes from right to left horizontally, with a fixed number of 40 bilateral eye movements per set.

In the IE phase, participants were asked to recall the traumatic event(s) or scene, with a focus on identifying the somatic, emotional, and physiological sensations that emerged. Each sensation was identified by the participant to the clinician, who instructed the participant to notice (focus on) the sensation while following the clinician’s oscillating hand to facilitate bilateral eye movements. This phase of identifying and processing sensations that emerged with IE was considered completed when the participant was able to re-imagine the entire traumatic experience with an acceptable (low) level of physiological reaction to the original distressing scene. In the IR phase, participants were instructed to visualize and imagine a new way to change or substitute the original negative traumatic experience with positive imagery, while also performing bilateral eye movements, as directed by the therapist. This technique of imaging new material to be added to or overwrite the original traumatic material is consistent with the process of memory reconsolidation (Lee, Nader, & Schiller, 2017). The IR phase was considered complete when the participant reported “seeing” (imagining) the memory in the new, preferred manner, as opposed to the original traumatic imagery. Additional information on the ART protocol has been previously published (Kip et al., 2012; Kip et al., 2014; Kip et al., 2013).

### Statistical Methods

Demographic and clinical characteristics of study participants by prior history of PTSD treatment status were compared using Fisher’s exact test for categorical variables and Student’s *t*-tests for continuous variables. Comparisons were made among the subjects who received and completed treatment with ART. Treatment outcomes were assessed based on mean score changes (pre-to-post ART treatment) on the PCL-M, CES-D, STICSA, BSI, and PSQI instruments, as well as by calculation of Cohen’s *d* effect size and corresponding 95% confidence intervals (Morris & DeShon, 2002). To examine potential differences in ART treatment response among groups for the first classification of prior PTSD treatment, analysis of covariance (ANCOVA) models were fit, including adjustment for baseline value of the evaluated outcome, PCL-M, PSQI, disability, antidepressant, antianxiety, and sleep medication. Moreover, the proportion of subjects who experienced a clinically and statistically meaningful reduction in symptoms of PTSD was defined using the established metric of ≥10-point reduction on the PCL-M (PTSD checklist) and compared by prior treatment history group by chi-square analysis. For the second classification of prior PTSD treatment, ANCOVA was again used with statistical adjustment for baseline value of the evaluated outcome, PCL-M, BSI, CES-D, disability, antidepressant, antianxiety, and medication use for sleep and pain. Statistical significance was set at a *p* <.05.

## Results

### Types of Prior History of PTSD Treatment (Classification #1)

#### Demographic Characteristics

Among 148 participants who enrolled and started treatment with ART, 106 (71.6%) completed treatment in a mean of 3.5 treatment sessions (Table 1). Of those who completed ART, 55 participants (51.9%) provided a 6-month follow-up data. The mean age of the study participants was 43.8 years, with 94.6% identifying as male, and 84.3% identifying as White. The study population was predominantly veterans (87.1%) with prior Army service (57.4%) and who had experienced high combat-related activity (81.8%), and 71.6% witnessed death or execution. Over half had trauma-related symptoms for 11 or more years (53.8%). Of note, 53.6% of participants in the first-line psychotherapy group were more likely on disability for PTSD or other mental health disorders, compared to the treatment-naïve group (15.6%), a statistically significant difference (*p* = 0.01).

**Table 1.** Demographic and Clinical Characteristics of Study Participants by Types of Prior History of PTSD Treatment (Classification #1)

| Characteristics | Types of Prior History of PTSD Treatment |  |  |  |  | *p*-Value |
| --- | --- | --- | --- | --- | --- | --- |
|  | All<br>(n=148) | Treatment-naïve (n=33) | Medication<br>(n=40) | Other<br>Psychotherapy<br>(n=42) | First-line<br>Psychotherapy<br>(n=33) |  |
| Demographic characteristics |  |  |  |  |  |  |
| Age in years, mean ± SD | 43.8±13.4 | 45.7±15.2 | 45.6±13.1 | 41.7±12.8 | 42.5±12.3 | 0.43 |
| Male gender, % | 94.6 | 93.9 | 90.0 | 100.0 | 93.9 | 0.25 |
| Race, % |  |  |  |  |  | 0.99 |
| White | 84.3 | 81.8 | 82.1 | 88.1 | 84.4 |  |
| Black | 13.0 | 15.2 | 15.4 | 9.5 | 12.5 |  |
| Other | 2.7 | 3.0 | 2.6 | 2.4 | 3.1 |  |
| Years of education, mean ± SD | 14.4±2.7 | 14.2±3.2 | 14.4±3.0 | 14.2±2.0 | 14.9±2.5 | 0.70 |
| Current military status, % |  |  |  |  |  | 0.64 |
| Active duty | 6.8 | 9.4 | 10.0 | 7.1 | 0.0 |  |
| Reservist | 6.1 | 9.4 | 5.0 | 4.8 | 6.1 |  |
| Discharged/veteran | 87.1 | 81.3 | 85.0 | 88.1 | 93.9 |  |
| Four or more overseas tour, % | 25.2 | 20.7 | 22.2 | 23.8 | 34.4 | 0.58 |
| Branch of service, % |  |  |  |  |  | 0.18 |
| Army | 57.4 | 42.4 | 62.5 | 64.3 | 57.6 |  |
| Navy | 12.8 | 24.2 | 10.0 | 7.1 | 12.1 |  |
| Marine Corps | 13.5 | 12.1 | 15.0 | 4.8 | 24.2 |  |
| Air force | 9.5 | 12.1 | 7.5 | 14.3 | 3.0 |  |
| National guard | 6.8 | 9.1 | 5.0 | 9.5 | 3.0 |  |
| Clinical characteristics |  |  |  |  |  |  |
| PCL-M (PTSD checklist), mean ± SD | 58.5±13.8 | 52.3±15.5 | 58.8±12.2 | 58.4±14.8 | 64.6±9.9 | 0.01* |
| Brief Symptom Inventory, mean ± SD | 28.4±13.7 | 24.5±15.3 | 28.6±12.4 | 27.1±13.6 | 32.3±14.1 | 0.18 |
| CES-D (depression), mean ± SD | 27.6±11.4 | 24.2±12.2 | 30.2±11.0 | 25.9±11.3 | 29.6±10.9 | 0.11 |
| STICSA (cognitive), mean ± SD | 24.2±7.2 | 22.8±8.1 | 25.3±7.3 | 22.9±6.8 | 25.9±6.7 | 0.20 |
| STICSA (somatic), mean ± SD | 20.3±5.9 | 20.2±6.7 | 20.7±6.3 | 19.7±4.8 | 20.5±6.1 | 0.90 |
| PSQI (sleep quality), mean ± SD | 13.4±3.9 | 12.0±3.8 | 13.9±3.8 | 12.6±4.0 | 15.0±3.3 | 0.01* |
| On disability for PTSD/MH, % | 36.1 | 15.6 | 35.0 | 37.5 | 56.3 | 0.01* |
| Previous trauma history, % |  |  |  |  |  |  |
| Sexual trauma | 10.1 | 12.1 | 17.5 | 2.4 | 9.1 | 0.15 |
| Physical assault/homicide of civilian | 16.9 | 18.2 | 17.5 | 11.9 | 21.2 | 0.75 |
| Combat-related activity | 81.8 | 73.3 | 73.7 | 90.5 | 87.9 | 0.11 |
| Witness death/execution | 71.6 | 66.7 | 67.5 | 73.8 | 78.8 | 0.64 |
| 5+ traumas/major injuries | 54.6 | 50.0 | 51.3 | 52.6 | 65.6 | 0.56 |
| Trauma for 11+ years, % | 53.8 | 65.6 | 55.0 | 53.7 | 40.6 | 0.26 |
| Current medications, % |  |  |  |  |  |  |
| Antidepressant | 46.0 | 12.1 | 77.5 | 38.1 | 51.5 | <.01* |
| Antianxiety | 21.6 | 3.0 | 32.5 | 11.9 | 39.4 | 0.01* |
| Sleep | 20.3 | 0.0 | 22.5 | 14.3 | 45.5 | <.01* |
| Antipsychotics | 12.2 | 3.0 | 12.5 | 9.5 | 24.2 | 0.06 |
| Pain | 33.8 | 15.2 | 37.5 | 42.9 | 36.4 | 0.07 |
| Treatment completion rate, % | 71.6 | 54.6 | 77.5 | 76.2 | 75.8 | 0.11 |
| Total ART sessions, mean ± SD | 3.5±1.4 | 3.0±1.5 | 3.6±1.3 | 3.5±1.4 | 3.7±1.5 | 0.19 |

PTSD: post-traumatic stress disorder; PCL-M: PTSD Checklist; CES-D: Center for Epidemiologic Depression Scale; STICSA: State-Trait Inventory for Cognitive and Somatic Anxiety; PSQI: Pittsburgh Sleep Quality Index; MH: mental health; SD: standard deviation; ART: Accelerated Resolution Therapy. \*Significant difference at $p < .05$ .

#### Clinical Presentation and Treatment Results

Study participants in the treatment-refractory groups (medication only, other psychotherapy, and first-line psychotherapy) had significantly higher mean baseline values on the PCL-M for PTSD and PSQI for sleep quality compared to the treatment-naïve group (*p* = 0.01, Table 1). The first-line psychotherapy (88.0%) and the treatment-naïve group (87.5%) had the highest ≥10-point reduction on the PCL-M post-treatment and at follow-up, respectively (Figure 1). As seen at the top of Table 2, all groups experienced significant pre-to-post within-group changes in PTSD symptoms: 15.4-point reduction in the medication group (effect size = 1.11), 24.1 points in the first-line psychotherapy group (effect size = 1.88), 19.2 points in the other psychotherapy group (effect size = 1.03), and 25.1 points in the treatment-naïve group (effect size = 1.48). Thus, participants with a prior history of first-line psychotherapy experienced substantial clinical benefit with ART, as did the treatment naïve group (*p* = 0.02 for between-group comparison). In assessing treatment-related changes in PTSD-related comorbidities, all groups showed similar modest-to-large improvements for symptoms of psychological distress, depression, anxiety, and sleep quality. At a 6-month follow-up, the largest sustained reductions in symptoms of PTSD occurred in the treatment naïve group (−28.3 points, effect size = 1.48) and the first-line psychotherapy group (−28.9 points, effect size = 1.53; Table 2, bottom).

**Figure 1.**
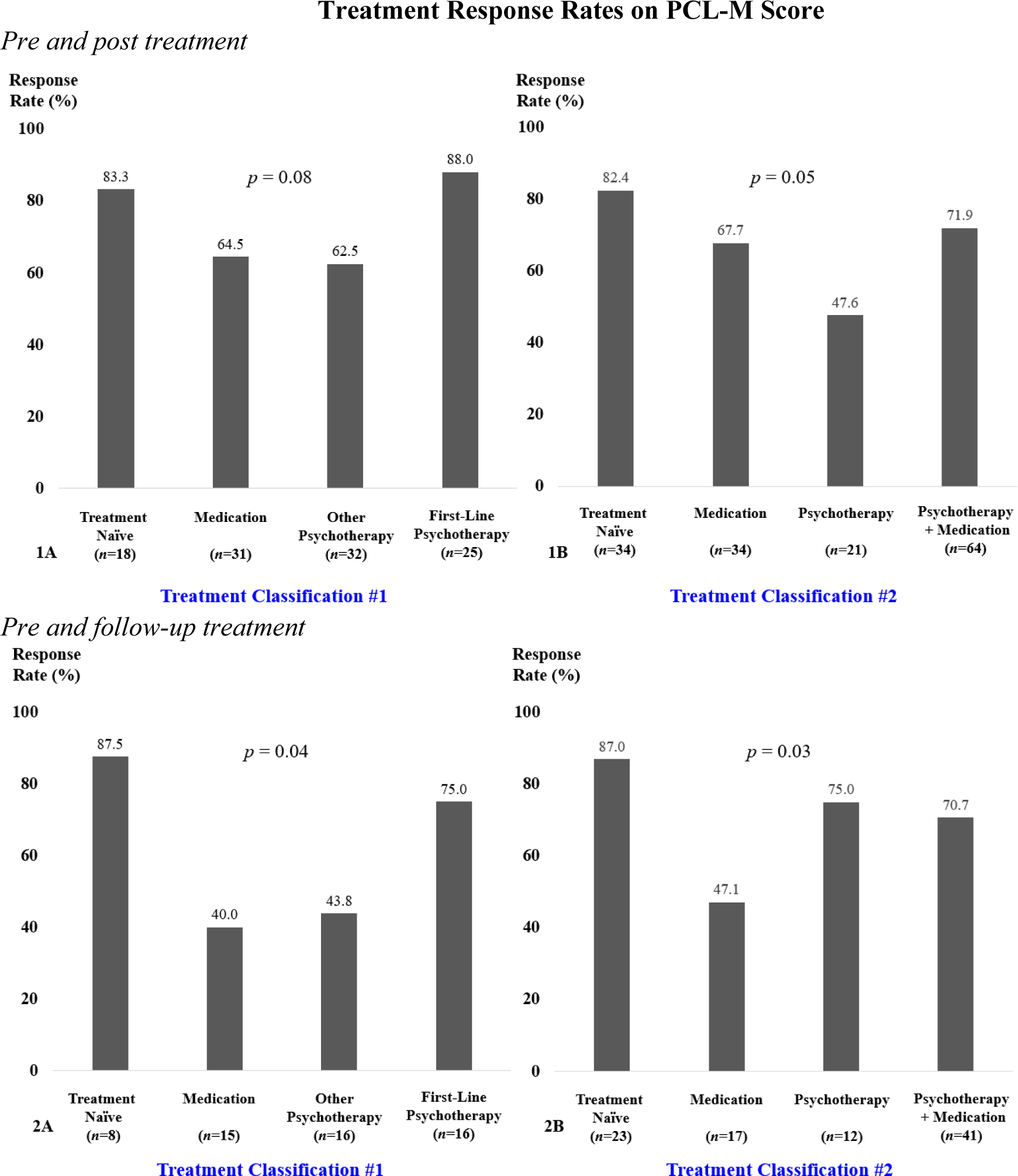
Histogram of treatment response rates defined as ≥10-point reduction in PTSD symptoms on the 17-item PCL-M (PTSD) checklist by classification of prior history of PTSD treatment. The top two figures (1A and 1B) present response rates among subjects pre and post treatment with ART. The bottom two figures (2A and 2B) present response rates among subjects pre and follow-up treatment with ART. ART: Accelerated Resolution Therapy; PTSD: post-traumatic stress disorder; PCL-M: PTSD Checklist. Classification #1 is types of prior history of PTSD treatment. Classification #2 is prior history of PTSD treatment

**Table 2.**
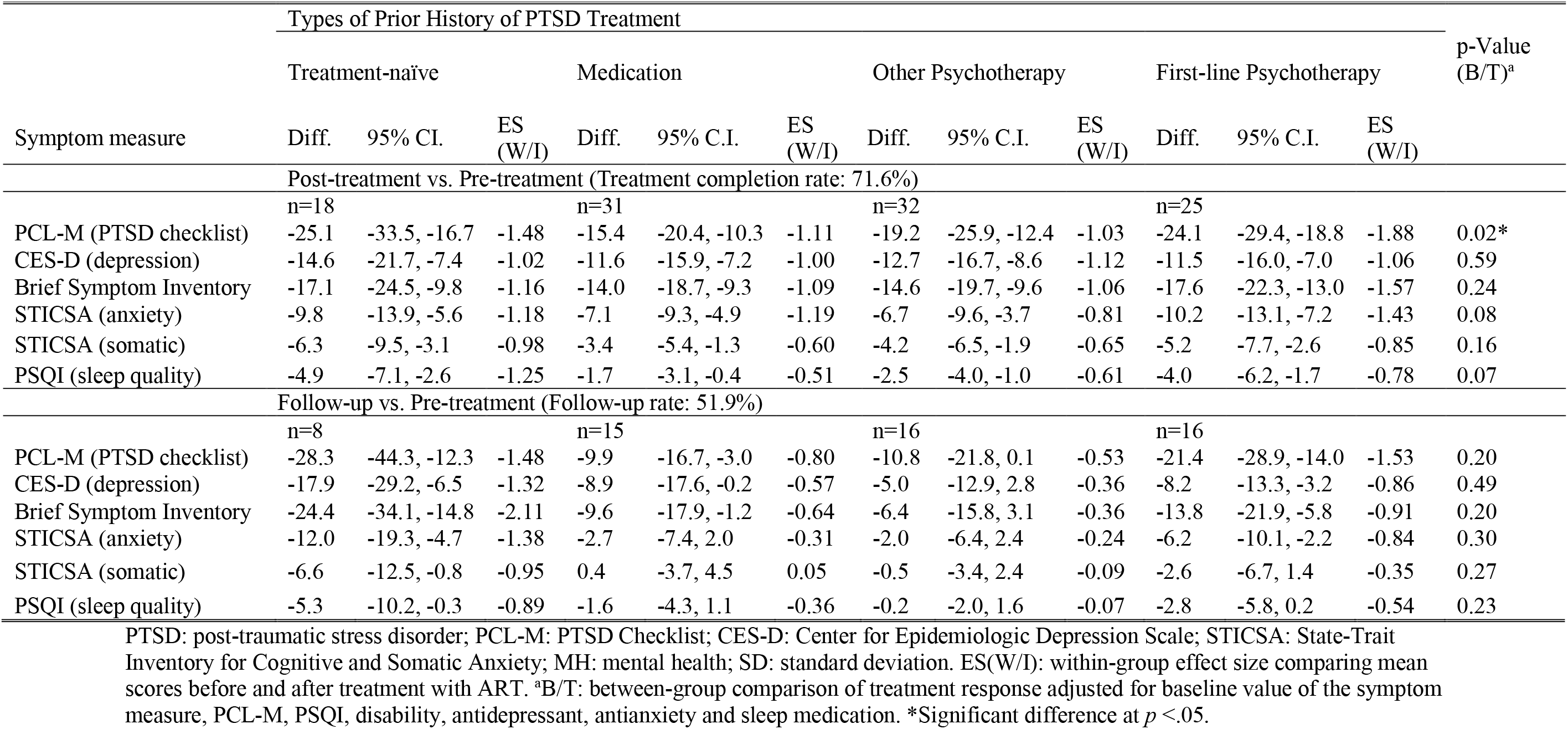
Symptom Treatment Response with Accelerated Resolution Therapy (ART) by Types of Prior History of PTSD Treatment (Classification #1)

### Prior History of PTSD Treatment (Classification #2)

#### Demographic Characteristics

Using the second definition of prior history of PTSD treatment, a total of 198 eligible military service members/veterans started treatment with ART, and 153 (77.3%) completed treatment in a mean of 3.5 treatment sessions (Table 3). Of the 153 subjects who completed treatment, 93 (60.8%) also provided 3- or 6-month follow-up data. The mean age of study subjects was 43.4±13.2 years, with 91.4% identifying as male, 84.2% identifying as White. Over half of the participants (52.8%) had trauma-related symptoms for more than 11 years. The majority of the study population were veterans (83.3%) with prior Army service (57.6%), had combat-related activity (84.4%), witnessed death or execution (72.1%), and had five or more traumas or major injuries (52.9%). Over half (53.3%) of study participants previously treated with medication and psychotherapy for PTSD were on disability for PTSD or other mental health disorders, compared to just 14.3% in the treatment-naïve group (*p* <.01).

**Table 3.** Demographic and Clinical Characteristics of Study Participants by Prior History of PTSD Treatment (Classification #2)

| Characteristics | Prior History of PTSD Treatment |  |  |  |  | p-Value |
| --- | --- | --- | --- | --- | --- | --- |
|  | All<br>(n=198) | Treatment-naïve (n=50) | Medication<br>(n=43) | Psychotherapy<br>(n=25) | Psychotherapy<br>+Medication<br>(n=80) |  |
| Demographic characteristics |  |  |  |  |  |  |
| Age in years, mean ± SD | 43.4±13.2 | 45.5±14.5 | 44.8±13.0 | 40.3±10.4 | 42.2±13.1 | 0.29 |
| Male gender, % | 91.4 | 94.0 | 90.7 | 100.0 | 87.5 | 0.22 |
| Race, % |  |  |  |  |  | 0.91 |
| White | 84.2 | 80.0 | 83.3 | 88.0 | 86.1 |  |
| Black | 11.6 | 14.0 | 14.3 | 8.0 | 11.4 |  |
| Other | 3.9 | 6.0 | 2.4 | 4.0 | 2.5 |  |
| Years of education, mean ± SD | 14.6±2.7 | 14.7±3.2 | 14.4±2.9 | 14.6±2.2 | 14.7±2.3 | 0.92 |
| Current military status, % |  |  |  |  |  | 0.44 |
| Active duty | 8.1 | 12.2 | 9.3 | 12.0 | 3.8 |  |
| Reservist | 8.1 | 12.2 | 4.7 | 8.0 | 7.5 |  |
| Discharged/veteran | 83.8 | 75.5 | 86.1 | 80.0 | 88.8 |  |
| Four or more overseas tour, % | 27.3 | 24.4 | 23.1 | 37.5 | 27.9 | 0.61 |
| Branch of service, % |  |  |  |  |  | 0.75 |
| Army | 57.6 | 44.0 | 60.5 | 60.0 | 63.8 |  |
| Navy | 14.7 | 22.0 | 14.0 | 8.0 | 12.5 |  |
| Marine Corps | 12.6 | 12.0 | 14.0 | 16.0 | 11.3 |  |
| Air force | 10.1 | 16.0 | 7.0 | 12.0 | 7.5 |  |
| National guard | 5.1 | 6.0 | 4.7 | 4.0 | 5.0 |  |
| Clinical characteristics |  |  |  |  |  |  |
| PCL-M (PTSD checklist), mean ± SD | 58.2±14.0 | 52.2±15.0 | 58.2±12.0 | 59.2±14.8 | 61.7±13.0 | 0.01* |
| Brief Symptom Inventory, mean ± SD | 27.9±14.4 | 23.2±14.8 | 27.4±12.9 | 27.0±15.1 | 31.3±14.1 | 0.03* |
| CES-D (depression), mean ± SD | 27.3±11.9 | 22.2±11.6 | 29.5±11.3 | 27.5±12.7 | 29.0±11.5 | 0.01* |
| STICSA (cognitive), mean ± SD | 24.0±7.4 | 21.8±7.7 | 24.5±7.6 | 25.0±7.1 | 24.7±7.1 | 0.17 |
| STICSA (somatic), mean ± SD | 20.1±6.1 | 19.5±6.2 | 20.2±6.3 | 20.0±6.7 | 20.4±5.8 | 0.89 |
| PSQI (sleep quality), mean ± SD | 13.2±4.0 | 11.7±4.1 | 13.8±3.7 | 13.6±3.8 | 13.6±4.0 | 0.05 |
| On disability for PTSD/MH, % | 37.1 | 14.3 | 34.9 | 36.0 | 53.3 | <.01* |
| Previous trauma history, % |  |  |  |  |  |  |
| Sexual trauma | 11.7 | 8.0 | 16.3 | 4.2 | 13.8 | 0.36 |
| Physical assault/homicide of civilian | 16.8 | 14.0 | 18.6 | 16.7 | 17.5 | 0.94 |
| Combat-related activity | 84.4 | 78.7 | 75.6 | 91.7 | 90.0 | 0.09 |
| Witness death/execution | 72.1 | 70.0 | 69.8 | 75.0 | 73.8 | 0.93 |
| 5+ traumas/major injuries | 52.9 | 51.0 | 52.4 | 56.5 | 53.3 | 0.98 |
| Trauma for 11+ years, % | 52.8 | 65.3 | 51.2 | 40.0 | 50.0 | 0.17 |
| Current medications, % |  |  |  |  |  |  |
| Antidepressant | 43.4 | 10.0 | 76.7 | 4.0 | 58.8 | <.01* |
| Antianxiety | 20.2 | 4.0 | 30.2 | 8.0 | 28.8 | <.01* |
| Sleep medication | 18.7 | 2.0 | 20.9 | 12.0 | 30.0 | <.01* |
| Antipsychotic | 9.6 | 2.0 | 11.6 | 12.0 | 12.5 | 0.21 |
| Pain reduction | 30.8 | 16.0 | 34.9 | 8.0 | 45.0 | <.01* |
| Treatment completion rate, % | 77.3 | 68.0 | 79.1 | 84.0 | 80.0 | 0.32 |
| Total ART sessions, mean ± SD | 3.5±1.3 | 3.2±1.4 | 3.7±1.3 | 3.4±1.3 | 3.7±1.3 | 0.25 |

PTSD: post-traumatic stress disorder; PCL-M: PTSD Checklist; BSI: Brief Symptom Inventory; CES-D: Center for Epidemiologic Depression Scale; STICSA: State-Trait Inventory for Cognitive and Somatic Anxiety; PSQI: Pittsburgh Sleep Quality Index; MH: mental health; SD: standard deviation; ART: Accelerated Resolution Therapy. \*Significant difference at $p < .05$ .

#### Clinical Presentation and Treatment Results

The treatment-refractory groups (medication only, psychotherapy only, and psychotherapy and medication) had higher baseline scores on the PCL-M for PTSD, BSI for psychological distress, and CES-D for depression compared to the treatment-naïve group (Table 3). The treatment-naïve group had the highest ≥10-point reduction on the PCL-M (post-treatment 82.4%, follow-up 87%), using the definition of statistical and clinically meaningful change (Figure 1). All treatment groups that completed treatment with ART experienced substantial pre-to-post reductions in PTSD symptoms (PCL-M): 16.5-point reduction in the medication-only group (effect size = 1.20), 15.5 points in the psychotherapy group (effect size = 0.86), 20.2 points in the medication and psychotherapy group (effect size = 1.31), and 20.6 points in the treatment-naïve group (effect size = 1.39; Table 4, top). The mean score reduction on the PCL-M was not statistically different between the four groups after adjusting for baseline value and other confounders (*p* = 0.19), indicating ART was approximately equally effective among all treatment-refractory groups and those with no prior treatment history. Similar reductions of comorbidities were observed in all groups after completing treatment with ART, including symptom measures of psychological distress, depression, anxiety, and sleep quality. The reductions in PTSD and its related symptoms generally persisted at 3- or 6-months follow-up (Table 4, bottom).

**Table 4.** Symptom Treatment Response with Accelerated Resolution Therapy (ART) by Prior History of PTSD Treatment (Classification #2)

| Symptom measure | Prior History of PTSD Treatment |  |  |  |  |  |  |  |  |  |  |  | p-Value (B/T)<sup>a</sup> |
| --- | --- | --- | --- | --- | --- | --- | --- | --- | --- | --- | --- | --- | --- |
|  | Treatment-naïve |  |  | Medication |  |  | Psychotherapy |  |  | Psychotherapy+ Medication |  |  |  |
|  | Diff. | 95% CI. | ES (W/I) | Diff. | 95% C.I. | ES (W/I) | Diff. | 95% C.I. | ES (W/I) | Diff. | 95% C.I. | ES (W/I) |  |
| Post-treatment vs. Pre-treatment (Treatment completion rate: 77.3%) |  |  |  |  |  |  |  |  |  |  |  |  |  |
|  | n=34 |  |  | n=34 |  |  | n=21 |  |  | n=64 |  |  |  |
| PCL-M (PTSD checklist) | -20.6 | -25.8, -15.4 | -1.39 | -16.5 | -21.3, -11.7 | -1.20 | -15.5 | -23.7, -7.3 | -0.86 | -20.2 | -24.0, -16.3 | -1.31 | 0.19 |
| CES-D (depression) | -12.5 | -16.6, -8.4 | -1.07 | -12.2 | -16.3, -8.1 | -1.05 | -12.9 | -17.6, -8.3 | -1.27 | -12.0 | -14.9, -9.1 | -1.03 | 0.56 |
| Brief Symptom Inventory | -15.4 | -20.3, -10.4 | -1.08 | -13.8 | -18.1, -9.4 | -1.10 | -13.6 | -19.0, -8.2 | -1.18 | -15.8 | -19.1, -12.6 | -1.20 | 0.40 |
| STICSA (anxiety) | -8.0 | -10.6, -5.4 | -1.09 | -6.9 | -9.0, -4.9 | -1.19 | -6.4 | -10.5, -2.4 | -0.72 | -8.1 | -9.8, -6.3 | -1.16 | 0.06 |
| STICSA (somatic) | -5.1 | -7.1, -3.1 | -0.87 | -3.4 | -5.3, -1.5 | -0.63 | -3.6 | -6.8, -0.3 | -0.50 | -4.2 | -5.7, -2.8 | -0.72 | 0.16 |
| PSQI (sleep quality) | -3.8 | -5.2, -2.4 | -1.06 | -1.8 | -3.1, -0.4 | -0.50 | -3.2 | -4.9, -1.5 | -0.90 | -2.6 | -3.9, -1.4 | -0.53 | 0.25 |
| Follow-up vs. Pre-treatment (Follow-up rate: 60.8%) |  |  |  |  |  |  |  |  |  |  |  |  |  |
|  | n=23 |  |  | n=17 |  |  | n=12 |  |  | n=41 |  |  |  |
| PCL-M (PTSD checklist) | -24.9 | -32.0, -17.7 | -1.53 | -11.1 | -17.3, -4.9 | -0.92 | -21.4 | -33.8, -8.9 | -1.09 | -17.5 | -22.4, -12.6 | -1.13 | 0.50 |
| CES-D (depression) | -13.3 | -18.5, -8.1 | -1.11 | -8.7 | -16.3, -1.1 | -0.59 | -12.2 | -20.6, -3.8 | -0.92 | -8.1 | -12.0, -4.1 | -0.65 | 0.78 |
| Brief Symptom Inventory | -19.7 | -25.9, -13.4 | -1.39 | -9.2 | -16.5, -1.8 | -0.64 | -17.3 | -26.5, -8.1 | -1.20 | -11.6 | -16.6, -6.6 | -0.74 | 0.83 |
| STICSA (anxiety) | -9.2 | -12.9, -5.6 | -1.12 | -2.5 | -6.6, -1.6 | -0.32 | -6.4 | -10.9, -1.9 | -0.91 | -4.7 | -7.0, -2.4 | -0.65 | 0.48 |
| STICSA (somatic) | -5.3 | -7.8, -2.8 | -0.94 | 0.3 | -3.3, 3.8 | 0.04 | -6.3 | -10.2, -2.5 | -1.04 | -1.8 | -3.6, 0.0 | -0.32 | 0.25 |
| PSQI (sleep quality) | -3.8 | -6.0, -1.6 | -0.79 | -2.0 | -4.6, 0.6 | -0.44 | -2.5 | -5.4, 0.4 | -0.57 | -1.5 | -3.0, 0.1 | -0.33 | 0.53 |

PTSD: post-traumatic stress disorder; PCL-M: PTSD Checklist; BSI: Brief Symptom Inventory; CES-D: Center for Epidemiologic Depression Scale; STICSA: State-Trait Inventory for Cognitive and Somatic Anxiety; MH: mental health; SD: standard deviation. ES(W/I): within-group effect size comparing mean scores before and after treatment with ART. <sup>a</sup>B/T: between-group comparison of treatment response adjusted for baseline value of the symptom measure, PCL-M, Brief symptom inventory, CES-D, disability, antidepressant, antianxiety, medication use for sleep and pain

## Discussion

Post-traumatic stress disorder is a debilitating psychiatric disorder, particularly among U.S. military service members and veterans. First-line treatments including Cognitive Processing Therapy (CPT) and Prolonged Exposure (PE) therapy, while moderately successful in randomized controlled trials within the civilian population, produce less-than-optimal outcomes for military-related PTSD (Steenkamp et al., 2020). Poor treatment retention, emotionally taxing interventions, and nonresponse rates point to a need for more efficacious psychotherapies for PTSD among servicemen and women (Najavits, 2015; Steenkamp et al., 2020).

This study examined immediate, short-term (three months), and long-term (six months) reductions in PTSD and related mental health symptoms for U.S. military service/veteran participants following completion of a new, innovative trauma-focused treatment modality – Accelerated Resolution Therapy (ART). The majority of participants had sought prior treatment, including psychotropic medications and first-line psychotherapies as recommended by guidelines from the VA/DoD (The Management of Posttraumatic Stress Disorder Work Group, 2017). Overall, ART was proven to be an effective psychotherapy in reducing post-traumatic stress symptoms and ancillary symptoms of psychological distress, depression, anxiety, and sleep quality after an average of only 3.5 ART sessions. Specifically, ART was shown to be equally effective at significantly reducing PTSD symptoms among groups with treatment-refractory PTSD or individuals with no prior PTSD treatment history. The treatment completion rate ranged from 70% to 80%, which appears to be higher than most first-line trauma-focused therapies (Foa et al., 2018; Najavits, 2015; Nidich et al., 2018; Rauch et al., 2019; Resick et al., 2017; Resick et al., 2015; Steenkamp et al., 2015; Steenkamp et al., 2020). Moreover, participants’ large symptom reductions persevered up to six months after ART, especially for individuals who had either never sought PTSD treatment in the past or had previously received a first-line psychotherapy treatment. In contrast, prior studies have shown that PTSD symptoms tend to persist among service members and veterans, particularly those with high combat experience and comorbid concerns like depression (Armenta et al., 2018). While not a direct randomized comparison, the present findings suggest that ART was equally if not more effective than most recommended treatments for PTSD among military service members and veterans, even those who have had combat experiences and comorbid mental health symptoms of depression and anxiety. The findings also suggest that ART may be more effective than medication, psychotherapy, or a combination of medication and psychotherapy for individuals who had never sought treatment before, even with high combat and comorbid depression.

Previous studies have attempted to isolate factors that contributed to treatment dropout, poor response rates, low help-seeking, and risks for persistent PTSD symptoms, with varying results (Armenta et al., 2018; Najavits, 2015; Xue et al., 2015). And while the reasons for unsuccessful prior treatment or lack of treatment-seeking in the past is not known about the current study’s sample population, common factors that have been identified in previous studies are combat experiences, PTSD symptom severity, poor sleep quality, and comorbid depression, anxiety, and other mental health disorders that can complicate treatment outcomes. The findings in the present study demonstrated that not only did participants’ PTSD symptoms improve significantly in a brief treatment period, so did their depression, sleep quality, anxiety, and overall psychological distress after completing ART. This suggests that ART may be particularly helpful for more severe and complex symptom constellations for U.S. military members and veterans.

### Limitations and Future Directions

The results of this study are a promising step toward identifying brief, affordable, less emotionally demanding, and successful treatment for persistent and treatment-refractory PTSD among U.S. military members and veterans; however, there are a few limitations. While the completion rate for ART was higher compared to most first-line trauma-focused psychotherapies, some participants did not complete treatment and did not complete the follow-up assessments. Some hypotheses are that symptoms remitted, and the client did not wish to return, or perhaps they were no longer clients of the VA at the time of follow-up. However, those who completed the treatment and provide follow-up data appeared to have higher rates of symptom reduction at follow-up than other first-line psychotherapies (Foa et al., 2018; Nidich et al., 2018; Rauch et al., 2019; Resick et al., 2017; Resick et al., 2015; Steenkamp et al., 2020). The present study’s sample population was representative of VA clients; however, these specific results cannot be generalized to other populations suffering from PTSD. Future research should be conducted on U.S. military service members and veterans in non-VA outpatient settings to compare these findings, as well as non-military civilian populations using the same assessments and follow-up timelines.

## Conclusions

Accelerated Resolution Therapy (ART) continues to show evidence as an effective treatment modality for PTSD (Kip et al., 2012; Kip et al., 2013; Kip et al., 2016) and related mental health concerns like depression (Kip et al., 2013), bereavement (Buck et al., 2020), and even pain (Kip et al., 2014). ART meets the VA/DoD Clinical Practice Guidelines (Department of Defense, 2017) criteria for first-line psychotherapies for the treatment of PTSD, as it is an individualized, manualized, trauma-focused psychotherapy (Kip & Diamond, 2018) and contains the most critical components of evidence-based trauma-focused treatment (Waits et al., 2017). Based on our results, not only should ART be considered as a first-line treatment for PTSD, but also this modality is to be given first consideration for military service members and veterans who are at high risk of treatment dropout and/or have treatment-refractory PTSD accompanied by other mental health symptoms. Similarly, ART should also be considered when minimal or no reductions in PTSD and related symptoms are observed following the initiation of PE, CPT, or similar treatments.

## Data Availability

Inquires related to the data that support the findings should be directed to the corresponding author. The data are not publicly available due to privacy and confidentiality requirements of the Institutional Review Board (IRB) at the University of South Florida (USF), the parent institution for the study. Requests for data access, if approved through the corresponding author and USF IRB, require a Data Authorization and Data Sharing Agreement between USF and the requesting institution.

## Acknowledgments

We thank the U.S. service members and veterans who participated in this research.

## Ethics statement

Ethical approval for this study was obtained by Institutional Review Board at University of South Florida (USF). All participants provided written informed consent.

